# Reflex single-gene non-invasive prenatal testing significantly increases the cost-effectiveness of carrier screening

**DOI:** 10.1101/2021.05.27.21256348

**Authors:** Shan Riku, Herman Hedriana, Jacqueline A. Carozza, Jennifer Hoskovec

**Affiliations:** BillionToOne, Inc., Menlo Park, CA, USA; University of California Davis Health, Sacramento, CA, USA

**Author notes:** To whom correspondence may be addressed: Jennifer Hoskovec, BillionToOne., Inc., 1035 O’Brien Drive, Menlo Park, CA, 94025.

**Keywords:** carrier screening, genetic disorders, pregnancy healthcare costs, decision-analytical model, cystic fibrosis, spinal muscular atrophy, thalassemia, sickle cell disease, genetic counseling, cell-free DNA

## Abstract

**Objective:** To evaluate the clinical and cost savings benefits of adoption of a carrier screen with reflex single-gene non-invasive prenatal test (sgNIPT) in prenatal care.

**Method:** A decision-analytic model was developed to compare carrier screen with reflex sgNIPT (maternal carrier status and fetal risk reported together) as first-line carrier screening to the traditional carrier screening workflow (positive maternal carrier screen followed by paternal screening to evaluate fetal risk). The model compared the clinical outcomes and cost effectiveness between the two screening methods. These results were used to simulate appropriate pricing for reflex sgNIPT.

**Results:** Reflex sgNIPT carrier screening detected 108 of 110 affected pregnancies per 100,000 births (98.5% sensitivity), whereas traditional carrier screening detected 46 of 110 affected pregnancies (41.5% sensitivity). The cost to identify one affected pregnancy was reduced by 62% in the reflex sgNIPT scenario compared to the traditional scenario. Adding together the testing cost savings and the savings from earlier clinical intervention made possible by reflex sgNIPT, the total cost savings was $37.6 million per 100,000 pregnancies. Based on these cost savings, we simulated appropriate reflex sgNIPT pricing range: if the cost to identify one affected pregnancy is the unit cost, carrier screening with reflex sgNIPT can be priced up to $1,859 per test (or $7,233 if sgNIPT is billed separately); if the cost per 100,000 pregnancies is the unit cost, carrier screening with sgNIPT can be priced up to $1,070 per test (or $2,336 if sgNIPT is billed separately).

**Conclusion:** Using the carrier screen with reflex sgNIPT as first-line screening improves the detection of affected fetuses by 2.4-fold and can save costs for the healthcare system. A real-life experience will be needed to assess the clinical utility and exact cost savings of carrier screen with reflex sgNIPT.

## Introduction

Autosomal recessive disorders represent a large disease burden worldwide.^1^ Manifestation of autosomal recessive disorders in the first 25 years of life is estimated to be 1.7 in 1,000^2^, and can be considerably higher in certain populations.^3^ Carrier screening is a genetic testing methodology that aims to identify individuals or couples who carry one variant allele within a gene and are at risk of having offspring with the associated genetic disorder.^4^ The American College of Obstetricians and Gynecologists (ACOG) recommends all patients who are considering pregnancy or currently pregnant be carrier screened for cystic fibrosis (CF), spinal muscular atrophy (SMA) and hemoglobinopathies.^4^ Most patients are screening after they become pregnant, with one 2019 study reporting that 436 out of 462 patients (94.4%) undergoing carrier screening were screened prenatally.^5^

The primary goal of carrier screening is to identify high-risk fetuses so that patients and providers can make informed decisions about undergoing diagnostic testing and considering prenatal and neonatal interventions. In the traditional carrier screen, pregnant patients are first carrier screened. While one in six pregnant patients is a carrier, only one in 900 newborns is affected with CF, SMA, or hemoglobinopathies, indicating that >99% of carrier pregnant patients carry unaffected fetuses. If the maternal carrier screen is positive, the next step is paternal carrier screening. If both parents are carriers, the fetus has a 1 in 4 risk of being affected and diagnostic testing is needed to determine disease status. However, the paternal carrier screen follow-up rate is low due to financial, societal, and logistical barriers.^6,7^ As a result of low paternal participation, traditional carrier screening fails to identify nearly 60% of affected pregnancies in the U.S.^5,7^, and it incurs costs associated with paternal screening and choice of prenatal and newborn interventions.

Single-gene non-invasive prenatal testing (sgNIPT) has the potential to overcome the challenges with traditional carrier screening workflow. A commercially available carrier screen with reflex sgNIPT was introduced into clinical use in 2019 in the U.S.^8^. This screen provides both maternal carrier status and fetal risk assessment from one maternal blood sample without the need for partner testing. Upon receipt of the maternal blood sample, carrier screening is performed for cystic fibrosis, spinal muscular atrophy, and alpha and beta hemoglobinopathies as recommended by ACOG^4^. Maternal carrier screening is performed via next generation sequencing on genomic DNA extracted from the maternal blood sample. If the pregnant patient is a carrier of any condition on the carrier screening panel, sgNIPT is then performed on the cell-free DNA extracted from the original sample as a reflex (see **Method S1**). Fetal risk assessment is provided as a personalized numerical risk and summarized as low risk (fetal risk <1/500), high risk (fetal risk >1/4), increased risk or decreased risk (fetal risk between 1/500-1/4) or no result. Within two weeks, the ordering health care provider receives both the maternal carrier result and the fetal risk which can be used to counsel the patient about options for diagnostic testing, particularly in the case of high-risk results (**Figure 1**). Reflex sgNIPT has an analytical sensitivity of >98% and specificity of >99.9%^8^.

**FIGURE 1.**
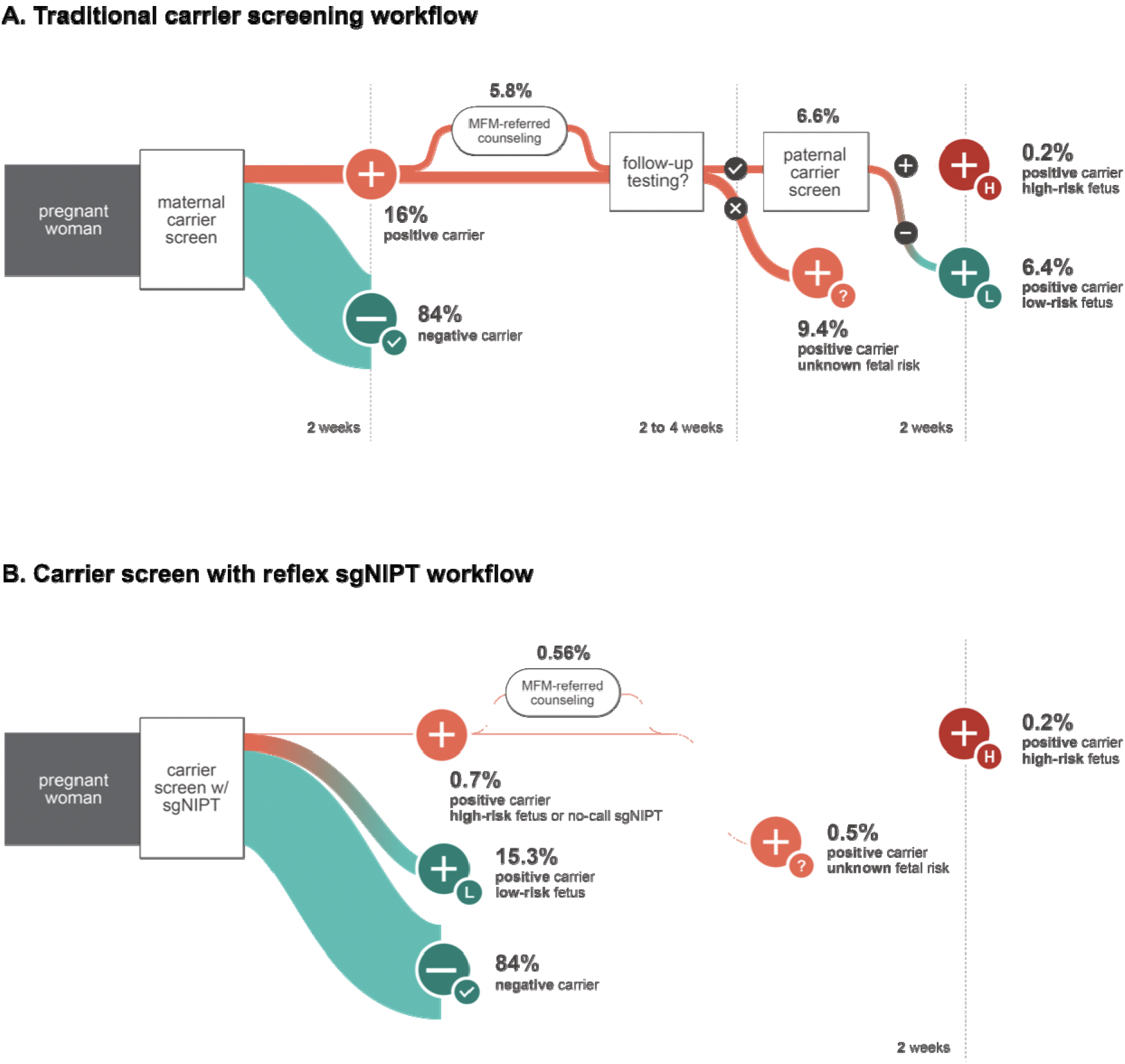
Schematic diagram of the decision tree model used for traditional carrier screen workflow (A) and reflex sgNIPT carrier screen workflow (B).

Here, we use a decision-analytical model to evaluate the clinical and cost saving impacts of adopting the carrier screen with reflex sgNIPT as a first-line screening test compared to the traditional carrier screen. We find that carrier screen with reflex sgNIPT detects more affected fetuses and saves $37.6 million per 100,000 pregnancies in cost to the healthcare system. Because the sgNIPT portion of the screen is not currently billed to payors or patients due to its recent commercial introduction, we use these cost savings results to explore appropriate pricing for the screen.

## Methods

### Decision-analytic model

A decision tree was developed to simulate the experience of pregnant patients who are unaware of their carrier status going through a traditional carrier screening process versus a reflex sgNIPT screening process (**Figure 1**). The model included patients with singleton pregnancies who elected maternal carrier screening. Patients undergoing preconception carrier screening and patients with multiple pregnancy were excluded. Since a small fraction of patients undergo maternal carrier screening prior to conception (e.g., 5.6% in a 2019 study^5^) and twin pregnancy rate is only 3.3% in the US^6^, this model includes ∼94% of patients undergoing maternal carrier screening. The traditional carrier screen base scenario was developed from the standard workflow typically carried out in obstetric clinics in the US. We did not account for the concurrent screening model where the mother and father of the pregnancy are tested simultaneously, as this model is not commonly used in obstetric clinics^9–12^. The carrier screen with reflex sgNIPT workflow (reflex sgNIPT scenario) was developed from the standard carrier screening workflow in obstetric clinics.

### Model inputs

Detailed assumptions and model inputs are provided in Supplementary Materials (**Method S2** and **Tables S1 and S2**). To simplify the model, several key assumptions were made as described below (basic workflow and test performance inputs are summarized in **Table 1**).

**TABLE 1.** A summary of clinical decision probabilities and test performance as part of the key assumptions for the decision-analytic model used in our analyses.

| Variables | Input Value |
| --- | --- |
| *Clinical decision probabilities (base scenario)* |  |
| MFM referral rate upon positive maternal carrier result | 36%* |
| Follow-up paternal carrier screening uptake | 41.5%<sup>5</sup> |
| *Clinical decision probabilities (UNITY scenario)* |  |
| MFM referral rate upon positive maternal carrier result | 80%** |
| *sgNIPT test performance* |  |
| Sensitivity | 98.5%<sup>8</sup> |
| Specificity | 99.9%<sup>8</sup> |
| sgNIPT no-call rate | 0.5%*** |

\* Based on internal OBGYN market research results. Four out of 11 OBGYNs (36%) interviewed refer patients to MFM clinic upon positive maternal carrier screen results.
\*\* Uptake for invasive diagnostic testing upon high-risk trisomy NIPT results are 73% for Trisomy 21, and 90% for Trisomy 18 and Trisomy 13.<sup>19</sup> Assuming a similar MFM referral rate for high-risk sgNIPT results to those for trisomy NIPT, we estimated the referral rate to be 80%.
\*\*\* Based on internal data
MFM, maternal-fetal medicine. sgNIPT, single-gene non-invasive prenatal testing; OBGYN, obstetrics and gynecology.

First, we only included the ACOG-recommended carrier screening workflow for cystic fibrosis (CFTR), spinal muscular atrophy (SMN1), sickle cell disease / beta hemoglobinopathies (HBB), and alpha thalassemia (HBA1/HBA2) in the analyses.

Second, the model assumed 100% carrier screening detection rates for both the base and reflex sgNIPT scenarios. Most laboratories in the U.S., including the carrier screen with reflex sgNIPT, offer NGS-based carrier screening which has 95-99+% detection rates for most genes.^13^

Third, the model focused on the comparison of workflows from the start of the carrier screening to the identification of high-risk fetuses, with both mother and father being identified as carriers in the base scenario and receiving a high-risk sgNIPT result in the reflex sgNIPT scenario. Due to the difference in how the tests are offered, uptake of the traditional carrier screening panels and the carrier screen with reflex sgNIPT panel among patients may vary. Further, due to the difference in reported fetal risk (25% fetal risk based on autosomal recessive inheritance with the traditional screening method, and 25-90% fetal risk based on internal criteria for carrier screen with reflex sgNIPT), uptake of diagnostic testing such as amniocentesis after the high-risk screening result may differ. However, given the insufficient data to fully simulate these differences, the uptake of diagnostic testing was excluded from the comparison model.

Fourth, because sgNIPT is not currently billed to payors or patients, we did not include the price of sgNIPT itself in the model. We assumed the same procedure cost inputs ($694 based on Fee schedules set by Center for Medicare and Medicaid Services; **Table S3**) for traditional carrier screen and the carrier screen portion of the carrier screen with reflex sgNIPT. We then explore appropriate pricing for carrier screen with sgNIPT using the results of the model.

### Sensitivity analysis

For sensitivity analysis, we used the total cost savings per 100,000 pregnancies as the model output. We then ran the model with the different values for each input individually (not considering interactions between the inputs for simplicity) to obtain the outputs and visualized the results with a tornado plot, arranged in order of largest impact on the model output. The estimates for low and high values are as follows: Paternal Follow-Up Rate: 20% and 80%; MFM Referral Rate: 10% and 70% (base scenario) or 40% and 100% (sgNIPT scenario) – the range is higher for sgNIPT since it provides a personalized fetal risk of up to 90% for high-risk cases; Diagnostic Testing Rate: 20% and 80% (both scenarios); Paternal Testing Cost: $694 (if there are no administrative costs) and $1,204 (if there are double administrative costs follow-up costs); MFM Cost: $818 and $1,618 (∼30% decrease and increase from original input); Diagnostic Testing Cost: $530 and $930 (∼30% decrease and increase from original input); % SMA Diagnosed Before Age 2 (Base): 40% and 80%; and % Eligible SMA Patients Choose Zolgensma: 50% and 100%.

## Results

### Clinical outcomes of base and reflex sgNIPT scenarios

The clinical outcome, detection of fetuses at high risk for common recessive disorders recommended by ACOG for prenatal screening (cystic fibrosis, spinal muscular atrophy, and hemoglobinopathies), was assessed for the base scenario (traditional workflow) and reflex sgNIPT scenario for 100,000 pregnancies (**Table 2**). Based on carrier rates in the U.S., 16,067 pregnant patients were screened to be positive carriers for one of the disorders for both scenarios (**Table S1**). The true number of affected fetuses is 110 out of 100,000 (0.11%), calculated from the incidence rates of each disorder.

**TABLE 2.** Clinical outcomes per 100,000 individuals screened in the base and reflex sgNIPT scenarios of our decision-analytical model and the differences between these two scenarios.

| Clinical outcomes | Base scenario | Reflex sgNIPT scenario | Difference |
| --- | --- | --- | --- |
| Number of positive maternal carriers | 16,067* | 16,067* | - |
| Number of paternal tests performed | 6,668 | - | (6,668) |
| Number of affected fetuses detected | 46 | 108 | 62 |
| Number of affected fetuses missed | 64 | 2 | (62) |
| Affected fetus detection rate (sensitivity) | 41.5%<sup>5</sup> | 98.5%<sup>8</sup> | 57.0% |

\* This number is based on the carrier frequency computed in **Table S1** per 100,000 individuals (100,000 × 16.1%).
sgNIPT, single-gene non-invasive prenatal testing.

Overall, reflex sgNIPT scenario detected far more affected fetuses than the base scenario. In the base scenario, 6,667 (41.5%) of the 16,067 positive carriers followed up with paternal testing based on previously published data.^5^ Therefore, 46 (41.5%) of 110 affected fetuses were detected through this scenario, and 64 affected fetuses (110-46 = 64; 58.5%) were missed due to the lack of paternal follow-up. In the reflex sgNIPT scenario, the maternal carrier status and fetal risk analysis are delivered to the patient together. Based on the 98.5% test sensitivity^8^, 108 out of 110 affected fetuses (110 * 98.5% = 108) were identified. Two out of 110 (1.5%) affected fetuses were missed through this workflow. Overall, there was a 2.4-fold increase in the detection rate of high-risk fetuses by carrier screen with reflex sgNIPT (108/110 identified) compared to the traditional workflow base scenario (46/110).

### Clinical utility and cost effectiveness of base and reflex sgNIPT scenarios

We then compared the clinical follow-up costs, including paternal testing and diagnostic testing, between base and reflex sgNIPT scenarios (**Method S3**). When the fetus was identified as high-risk in the base scenario (fetal risk is 25% if both the maternal and paternal carrier screens are positive) or reflex sgNIPT scenario (fetal risk ranges from 25% to 90%), we considered the affected pregnancy detected regardless of the diagnostic testing uptake.

The reflex sgNIPT scenario reduced the need for paternal screening, thus significantly lowering follow-up costs per 100,000 pregnancies. Follow-up testing in the base scenario costs $9.3M per 100,000 pregnancies, the sum of follow-up counseling and paternal screening ($9.2M) and diagnostic testing cost for high-risk fetuses ($0.1M) (**Table 3**). In contrast, follow-up testing in the reflex sgNIPT scenario costs $1.0M per 100,000 pregnancies, the sum of follow-up counseling ($0.7M) and diagnostic testing ($0.3M) (**Table 3**). Therefore, reflex sgNIPT saved $8.3M in follow-up costs per 100,000 pregnancies by reducing need for paternal screening. Internal quality assurance data supports that patients who receive low-risk sgNIPT results do not typically seek paternal screening.

**TABLE 3.** Total cost savings from reflex sgNIPT follow-up testing costs per 100,000 pregnancies.

| Item | Base Scenario | Reflex sgNIPT Scenario | Follow-up Cost Savings |
| --- | --- | --- | --- |
| **Follow-up cost with positive results (\$MM)** | **\$9.2** | **\$0.7** | **\$8.5** |
| *% carrier results that need follow-up (%) - ACOG panel* | 16.1% | - |  |
| *% follow-up needed for UNITY panel* | - | 1% |  |
| *% patients completing paternal testing* | 42% | - |  |
| *Paternal Test follow-up cost per case (Test & Admin)* | \$944 | - | |
| *% patients referred to MFM* | 36% | 80% |  |
| *Additional MFM follow-up cost per case* | \$1,236 | \$1,236 | |
| **Follow-up diagnostic testing cost (\$MM)** | **\$0.1** | **\$0.3** | **-\$0.2** |
| *% of positive paternal result* | 2.7% | - |  |
| *% diagnostic testing uptake* | 50% | 50% |  |
| *Cost of diagnostic testing* | \$730 | \$730 | |
| **Total follow-up cost (\$MM)** | **\$9.3** | **\$1.0** | **\$8.3** |

MFM, maternal-fetal medicine; SMA, spinal muscular atrophy.

We also calculated that the cost to identify one affected fetus is several times lower in the reflex sgNIPT scenario compared to the base scenario. To calculate the cost to identify one affected fetus, we first summed the total cost per 100,00 pregnancies (including the maternal screening cost of $694 per pregnancy) for each scenario and then divided by the number of fetuses detected. The base scenario identified 46 out of 110 affected fetuses (**Table 2**) at a cost of $78.7M, resulting in a cost of $1.73M per affected fetus (**Figure 2**). The reflex sgNIPT scenario identified 108 out of 110 affected pregnancies (**Table 2**) at a cost of $70.4M, resulting in a cost of $0.65M per affected fetus (**Figure 2**). Not including the cost of performing sgNIPT itself since it is not yet billed, the reflex sgNIPT scenario reduces the cost to identify one affected pregnancy by 62% (2.6-fold) from the base scenario (**Figure 2**) by identifying more affected fetuses and lowering the paternal screening cost. Of note, in either scenario the cost to screen one affected fetus is lower than the lifetime treatment cost (the weighted average lifetime treatment cost for the disorders screened is estimated to be $2.6M per patient; details provided in **Method S4** and **Table S6**).

**FIGURE 2.**
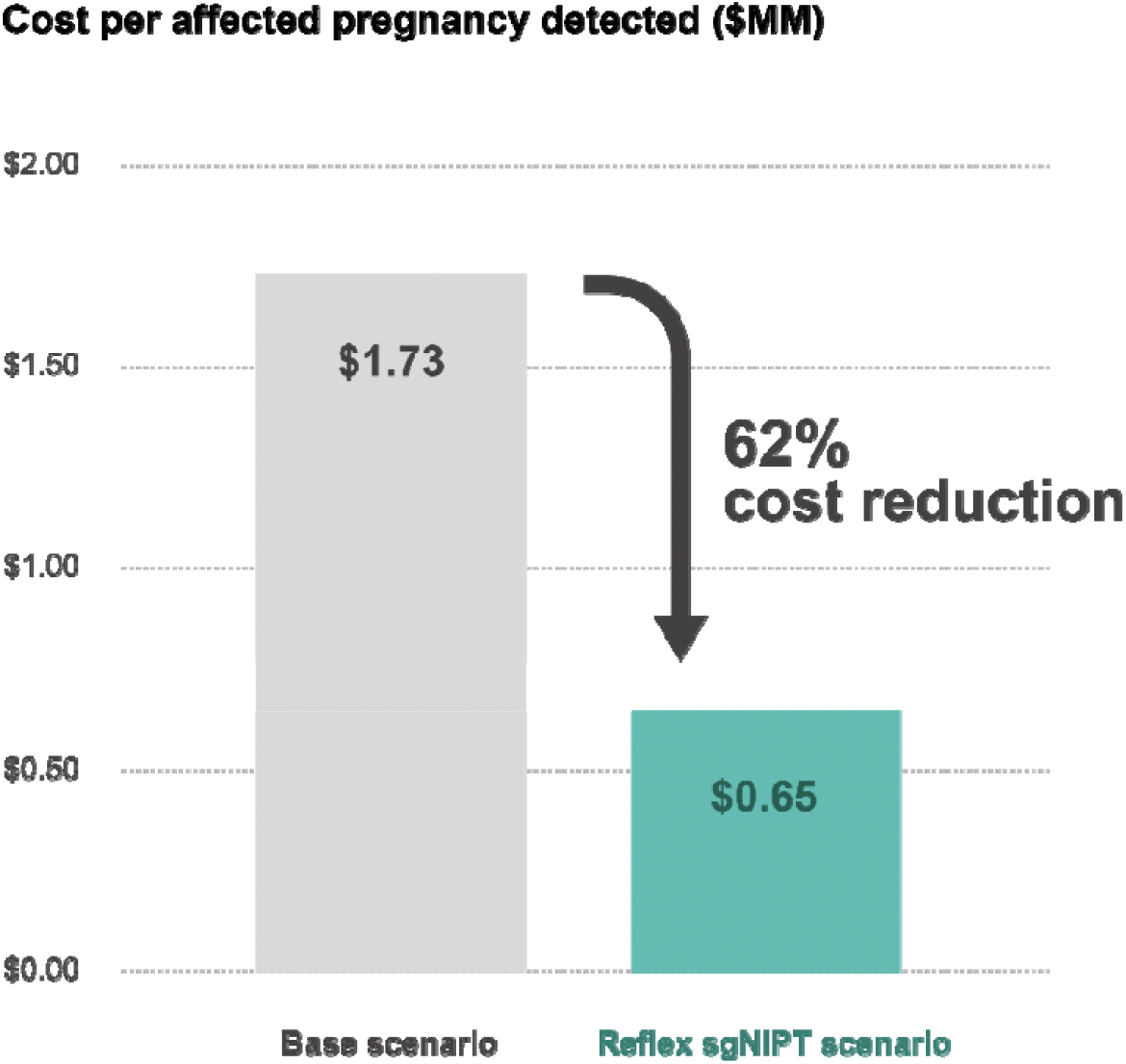
Comparison of cost effectiveness between base and reflex sgNIPT scenarios of detecting one affected pregnancy. Cost was calculated by dividing the total screening cost by the number of affected pregnancies detected.

### Clinical impact and healthcare cost saving from SMA treatment selection

Early identification of high-risk fetuses allows timely prenatal and neonatal interventions for clinicians and patients (**Table 4**), which in turn can lead to more effective management and potential cost savings. In particular, SMA has several treatment options, including Zolgensma and Spinraza. These therapies are most effective when delivered neonatally prior to onset of symptoms. Moreover, the single-dose gene therapy Zolgensma ($2.1M per dose) is more cost-effective than the lifetime medication Spinraza ($0.75M for initial dose and $0.46M per year afterward) (**Table S4, Table S5**). However, Zolgensma must be administered to neonates before two years of age. Since SMA is not included in newborn screening panels in all U.S. states (twelve states do not test for SMA as of June 2021), prenatal SMA diagnosis is essential for affected families to have access to early, clinically effective, and cost effective treatment options.^14^ The reflex sgNIPT scenario identifies more SMA-affected fetuses than the base scenario, resulting in more newborns eligible for Zolgensma. We estimated the total cost savings from more access to Zolgensma in the reflex sgNIPT scenario to be $29.3M per 100,000 pregnancies (**Table 5, Method S4**).

**TABLE 4.** Available prenatal and neonatal interventions for conditions screened by the reflex sgNIPT workflow.

| Condition | Disease description | Prenatal and neonatal interventions |
| --- | --- | --- |
| Spinal Muscular Atrophy (SMA) | • severe muscle weakness and wasting<br>• loss of motor skills<br>• breathing problems<br>• life expectancy for Type 1 (most severe) if untreated = 2 years<br>• most common genetic cause of infant death | • Spinraza, Zolgensma, and Evrysdi (genetic treatments administered after birth) increase survival and improve symptoms<br>• early treatment before onset of symptoms is crucial for effectiveness |
| Cystic Fibrosis (CF) | • breathing problems,<br>• severe lung damage<br>• serious digestive problems<br>• symptoms worsen over time<br>• average life expectancy = mid-thirties. | • medication and therapies improve symptoms and quality of life<br>• early diagnosis and careful lifelong management may increase life expectancy |
| Sickle Cell Disease / Beta Hemoglobinopathies | • fewer healthy red blood cells<br>• fatigue and pain<br>• complications such as poor growth, infection, heart problems, and stroke<br>• shortened life expectancy | • medications, regular blood transfusions, and bone marrow/stem cell transplants manage disease<br>• gene therapy may be available for some patients<br>• prenatal screening helps identify pregnancies at risk that may benefit from cord blood banking at birth |
| Alpha Thalassemia | • fewer healthy red blood cells<br>• alpha thalassemia major (most severe) results in death before birth or shortly after in untreated | • prenatal care essential to reduce risk of maternal complications for pregnancies with alpha thalassemia major<br>• prenatal treatment may increase survival likelihood |

sgNIPT, single-gene non-invasive prenatal testing; SMA, spinal muscular atrophy; CF, cystic fibrosis.

**TABLE 5.** Total cost savings from SMA treatment selection per 100,000 pregnancies.

| Item | Base<br>Scenario | Reflex<br>sgNIPT<br>Scenario | Cost<br>Savings |
| --- | --- | --- | --- |
| Number of SMA fetuses per 100,000 births<br>(prevalence) | 9 | 9 | - |
| % SMA patients diagnosed before age 2<br>(prenatally or postnatally) | 70% | 99% | - |
| % eligible patients (= diagnosed under age 2)<br>choosing Zolgensma as treatment | 90% | 90% | - |
| Per SMA patient lifetime treatment cost (\$MM) | **\$6.8** | **\$3.5** | - |
| **Total SMA treatment cost for 100,000 births**<br>**(\$MM)** | **\$60.3** | **\$31.1** | **\$29.3** |

SMA, spinal muscular atrophy.

### Overall cost savings from adoption of carrier screen with reflex sgNIPT

Together, we estimate that the cost savings in the reflex sgNIPT scenario (not including the cost of sgNIPT itself) is $37.6M per 100,000 pregnancies, which is the sum of follow-up cost savings ($8.2M) (**Table 2**) and more cost-efficient clinical intervention for fetuses affected with SMA ($29.3M) (**Table 5**).

We analyzed the sensitivity of the model output, the cost savings per 100,000 pregnancies, towards model inputs and found that sgNIPT saves costs compared to the base scenario over a wide range of input values (**Figure 3**). The inputs can be grouped into two categories: those that affect follow-up costs, and those that affect SMA treatment costs. The cost savings per 100,000 births is most sensitive to inputs that affect SMA treatment costs, including the base scenario SMA diagnosis rate before age two and the number of eligible SMA patients who chose Zolgensma. For example, a low base diagnosis rate (40%) results in relatively higher cost savings (up to $68.1M) and a high base diagnosis rate (80%) results in relatively lower cost savings ($27.4M). Additionally, the model is also sensitive to the percent of eligible SMA patients who choose Zolgensma over Spinraza. These two inputs are difficult to know with confidence because several U.S. states do not offer SMA newborn screening or are in the process of implementing it,^14^ and data on long-term efficacy and patient choices is not yet available^15^. Therefore, the exact cost savings for the sgNIPT scenario will vary within the U.S. depending on local SMA diagnosis rate before age two and availability of treatments.

**FIGURE 3.**
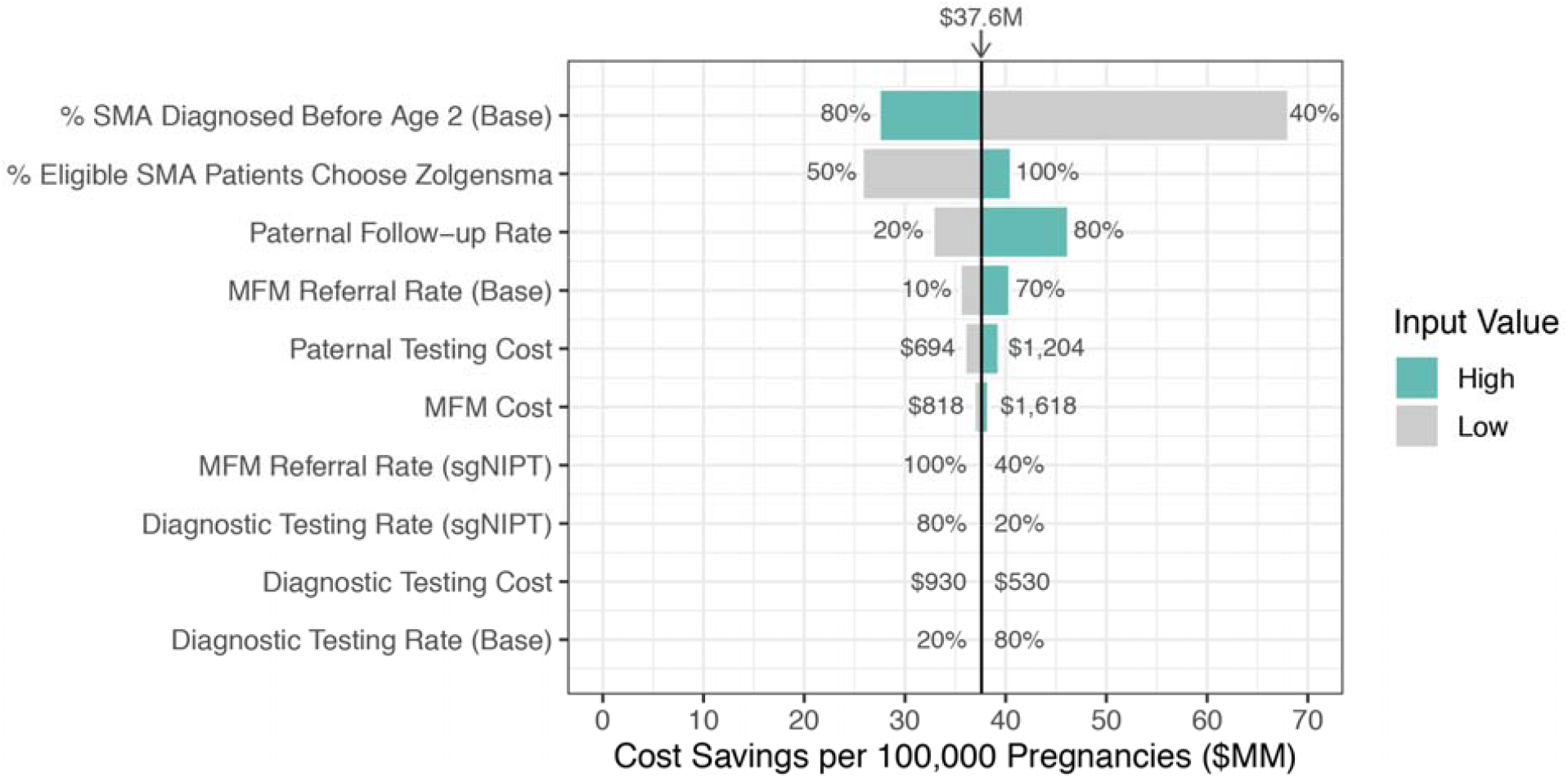
Sensitivity to inputs of reflex sgNIPT scenario cost savings per 100,000 pregnancies. The cost savings were calculated using a high and low value for each indicated input. The input value range is annotated next to each bar. The input values used in the model to yield $37.6M in cost savings are as follows: % SMA Diagnosed Before Age 2 (Base) = 70%; % Eligible SMA Patients Choose Zolgensma = 70%; Paternal Follow-Up Rate = 41.5%; MFM Referral Rate (Base) = 36%; Paternal Testing Cost = $944; MFM Cost = $1,236; MFM Referral Rate (sgNIPT) = 80%; Diagnostic Testing Rate (sgNIPT) = 50%; Diagnostic Testing Cost = $730; Diagnostic Testing Rate (Base) = 50%.

Out of the inputs that impact follow-up costs, the model is most sensitive to the base scenario paternal testing follow-up rate and relatively insensitive to other follow-up testing costs and referral rates. If paternal follow-up is low (20%), the cost savings for sgNIPT decrease to $32.8M per 100,000 pregnancies because fewer follow-up tests are needed (**Figure 3**). If paternal follow-up is high (80%), cost savings for sgNIPT increase to $46.2M per 100,000 pregnancies (**Figure 3**). Note that the effect is reversed for the cost to detected one affected fetus. If paternal follow-up is low (20%), fewer affected fetuses are identified which increases the cost to detected one affected fetus from $1.73M to $3.36M (**Table S7**). If paternal follow-up is high (80%), more affected fetuses are identified which decreases the cost to detected one affected fetus from $1.73M to $0.99M (**Table S7**). In both cases, sgNIPT cost to identify one affected fetus ($0.65M) remains the lowest.

### Additional considerations for non-U.S. healthcare systems

While the termination of pregnancy (TOP) is another available intervention available after early identification of affected fetuses, we did not include it as part of our U.S.-focused cost savings calculation. However, TOP is commonly considered outside the U.S. in nationwide implementations of prenatal screening programs. The disorders in our panel significantly impact quality of life and shorten lifespan even with current availability of treatments (**Table 4**). On average, lifetime treatment cost is $2.62M per patient (**Table S6**). We estimate that implementation of reflex sgNIPT could reduce lifetime treatment costs by $145.4M per 100,000 pregnancies due to potential TOP (**Table S8, Method S5**). The lifetime treatment cost inputs in the this model are derived from U.S. healthcare costs, and may need to be adjusted for other countries.

### Pricing simulations for carrier screen with reflex sgNIPT

The cost savings analysis above does not include the price of reflex sgNIPT since it is not yet billed to payers or patients. Therefore, the price of the reflex sgNIPT itself will decrease total cost savings. We used the results from our cost savings analysis to perform two simulations of appropriate pricing for carrier screen with reflex sgNIPT based on the value provided and identify price ranges that will save costs for the healthcare system overall (**Method S6**).

In the first simulation, we take the unit cost to be the cost to identify one affected fetus. The cost savings value is $1.08M per affected fetus ($1.73M in the base scenario - $0.65M in the reflex sgNIPT scenario). If we pass 100% of the cost savings value to the price of carrier screen with reflex sgNIPT, then the parity price is $1,859 (**Figure 4A, Method S6**). If reflex sgNIPT portion was priced separately from the carrier screen portion the parity price is $7,233, several-fold higher than the combined carrier screen with reflex sgNIPT price since the cost is divided only among carriers reflexed to sgNIPT instead of among all pregnancies carrier screened.

**FIGURE 4.**
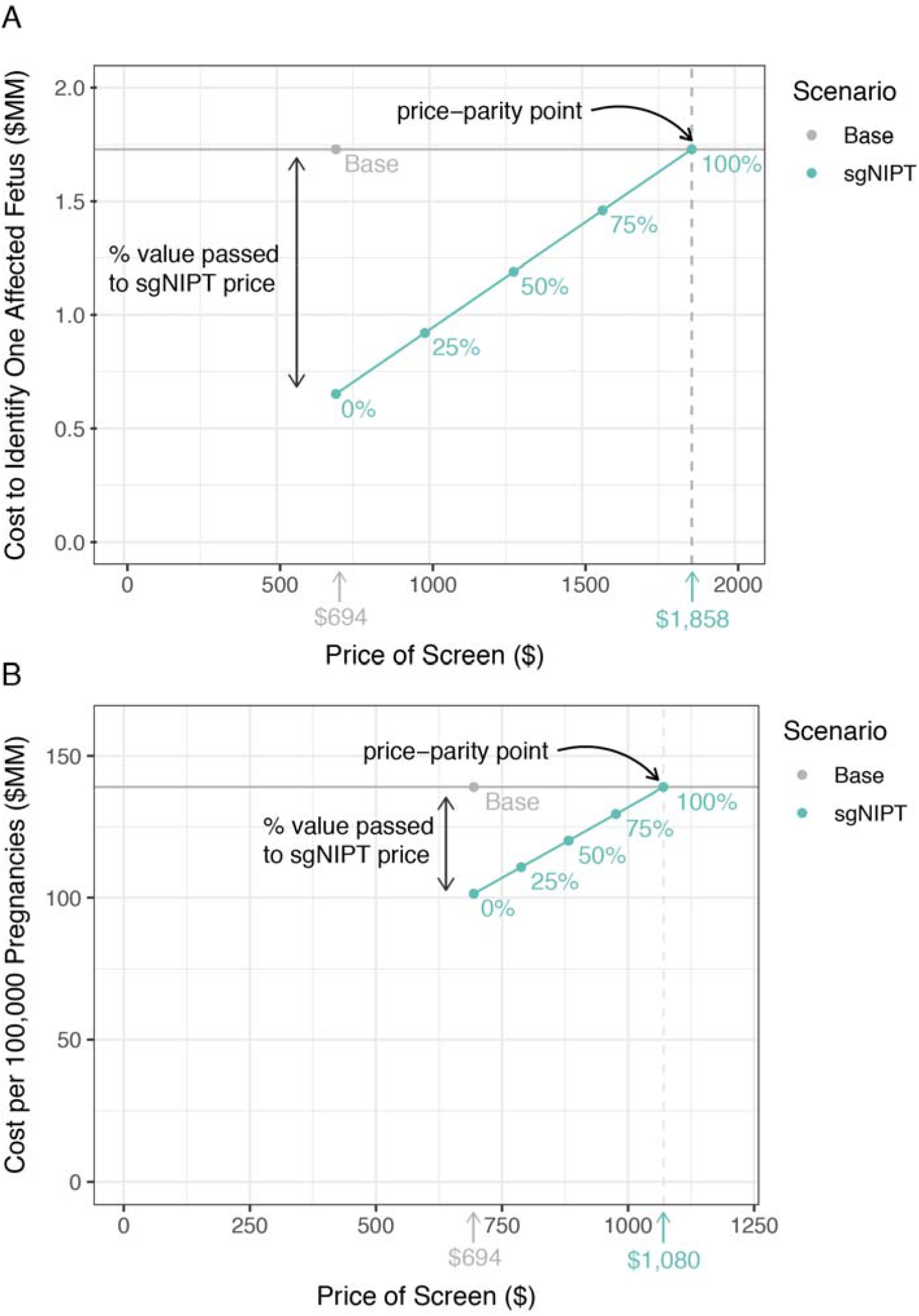
Pricing simulation for carrier screen with reflex sgNIPT using two different unit costs to determine the price-parity point with base scenario costs: (A) the cost to identify one affected fetus and (B) the cost per 100,000 pregnancies. Percentages refer to the percent of value (determined by the base costs) passed to the reflex sgNIPT costs.

In the second simulation, we take the unit cost to be the cost of carrier screening and downstream treatment per 100,000 pregnancies. As previously calculated, the cost savings value is $37.6M per 100,000 pregnancies. If we pass 100% of the cost savings value to the price of carrier screen with reflex sgNIPT, then the parity price is $1,070 (**Figure 4B, Method S6**). If reflex sgNIPT portion was priced separately from the carrier screen portion, the parity price is $2,336.

## Discussion

Results from our decision-analytic model analyses show that compared to the traditional sequential carrier screen workflow, the carrier screen with reflex sgNIPT workflow improves clinical outcome (detection of affected fetuses) and saves costs by eliminating paternal carrier screening and enabling more cost-effective SMA treatment interventions. Furthermore, we identified carrier screen with reflex sgNIPT pricing that would preserve cost savings for the healthcare system.

Carrier screen with reflex sgNIPT detects more affected pregnancies with less burden on the healthcare system. Although the traditional workflow is effective for detecting carriers of autosomal recessive conditions, the detection rate of high-risk fetuses is limited to 41.5%, as the lack of paternal follow-up testing is significant and leaves >58% of carrier pregnant patients without a fully informative risk assessment for their pregnancy.^5,7^ In contrast, the reflex sgNIPT carrier screen provides patients and physicians with a personalized risk for the fetus, allowing for more informed counseling, decision-making, and patient autonomy regarding follow-up testing and interventions. Additionally, the reduced need for paternal follow-up testing may significantly lower the burden on the physician, clinical staff, and patient, given the logistics involved in counseling about and arranging paternal testing.

Second, the reflex sgNIPT workflow reduces the time from blood draw to identification of high-risk fetuses by up to eight weeks compared to the traditional workflow. Early identification of high-risk fetuses will improve pregnancy and delivery management, patient counseling and education, and access to early intervention, therapeutics, research studies, and clinical trials. With pregnancies affected with alpha thalassemia, for example, only patients at 18 to 26 weeks gestational age are eligible for the in-utero hematopoietic stem cell transplantation clinical study.^16^ Faster turnaround time with reflex sgNIPT increases the likelihood of meeting the eligibility criteria and receiving treatment. Several treatment options, for example gene therapies for SMA, are most effective when implemented prior to the onset of symptoms. Uptake of genetic counseling and patient education after a positive newborn screen is low for some conditions.^14^ Early prenatal identification of high-risk fetuses makes it possible to connect the affected families with medical professionals who can educate and support them regarding options for diagnosis, treatment, and interventions.

The findings reported herein should be viewed considering several limitations. First, we recognize that paternal follow-up rates may vary between populations, within the U.S. or among different countries. The model can be refined if additional data on paternal follow-up rates are available. Second, clinical performance metrics in this report could change when data from a larger clinical study on the carrier screen with reflex sgNIPT become available. Third, since paternal carrier screening likely remains useful for future pregnancies of identified carriers or may be required by some laboratories as a control during diagnostic testing, the reflex sgNIPT workflow may not completely eliminate the cost of paternal testing, although it likely would reduce the urgency and frequency of the test. Fourth, cost savings from prenatal and neonatal interventions were calculated based on limited available literature, as many of these interventions are recent and rapidly evolving. While some of the data used in the models is dated, we chose the available data that was most applicable to the carrier screen clinical scenario. In particular, long-term data about extended life expectancy and outcomes for SMA patients treated with Spinraza or Zolgensma are still lacking, and the phenotypic severity is difficult to predict^15^. In addition, SMA newborn screening is rapidly changing and varies widely between countries, which would impact the cost savings amount.^14^ Furthermore, we expect that treatments will improve rapidly in clinical and cost effectiveness (e.g., stem cell transplantation for hemoglobinopathies^16–18^). Despite the above limitations, the current study provides a comprehensive assessment, using available data, on the clinical outcomes and cost effectiveness when using carrier screen with reflex sgNIPT as a first-line screening method versus the traditional sequential screening workflow. Furthermore, the model inputs and calculations can be updated (**Supplementary File 1**) to accommodate new or situation-specific data.

## Conclusion

Carrier screening with reflex sgNIPT improves clinical outcome by detecting fetuses at high risk for autosomal recessive disorders with higher sensitivity, giving patients and providers access to additional prenatal and neonatal intervention options. Furthermore, carrier screening with reflex sgNIPT can lead to significant cost savings to the healthcare system. While cost savings depends on the carrier screen with sgNIPT pricing (the current commercial test does not bill it), we used our model to explore pricing that saves costs for the system. If we define the unit cost as the cost to identify one affected fetus, a price less than $1,859 per carrier screen with reflex sgNIPT (or $7,233 if sgNIPT is billed separately) will save cost for the system. If we define the unit cost as the cost to screen 100,000 births, a price less than $1,070 per carrier screen with reflex sgNIPT (or $2,336 if sgNIPT is billed separately) will save cost for the system. Follow-up clinical studies are needed to assess the real-life clinical utility and cost savings of carrier screen with reflex sgNIPT.

## Data Availability

The data that support the findings of this study are available from the corresponding author upon request.

## Transparency

### Author Contributions

S.R. built the decision-analytical model. S.R. and J.A.C analyzed data. All authors wrote and edited the manuscript.

## Acknowledgements

We thank Dr. Oguzhan Atay, Dr. John ten Bosch, and Carrie McGehee for providing valuable inputs to the decision-analytic model, and Dr. Rong Mao for providing editorial support for this manuscript.

