## Supplementary Information for "Reflex single-gene non-invasive prenatal testing significantly increases the cost-effectiveness of carrier screening"

### SUPPLEMENTARY MATERIALS

#### Method S1: Overview of sgNIPT fetal risk assessment

Maternal carrier screening is performed by NGS on the genomic DNA extracted from a maternal blood sample. sgNIPT is performed on the cell-free DNA extracted from the original sample as a reflex. Pathogenic paternal alleles for the HBB, CFTR and HBA genes are detected by amplification and sequencing of select regions of each of gene. Maternal inheritance of a particular variant is assessed via relative mutation dosage, an examination of the variant allele fraction combined with cfDNA molecule counts using BillionToOne Quantitative Counting Technology (QCT)[8]. Fetal risk assessment is then calculated using data from the paternal allele assay, relative mutation dosage analysis, fetal fraction calculation, and a priori disease risk.

**Method S2: Demographic and Carrier Frequency Inputs**

***Ethnicity breakdown***

For the ethnicity breakdown of the women undergoing carrier screening, we used the 2018 report on US demographics for the Millennials generation (**Table S1**).[1]

***Carrier Frequency***

Carrier frequency for the conditions screened for are based on standard carrier frequency numbers found in literature (**Table S1**).

***Prevalence of at-risk pregnancies and affected fetus***

We defined "at-risk pregnancies" as the pregnancy between couples who are both carriers for the same disease, as there is a 25% chance for the baby to be affected. To estimate the incidence of at-risk pregnancy, we assumed that all couples conceived within their ethnicity group. For example, the prevalence of at-risk pregnancy for cystic fibrosis within Northern European ethnicity would be 4.0% × 4.0% = 0.16%. The combined prevalence of at-risk pregnancies for all conditions and ethnicities are estimated to be 2.51%.

To estimate the number of affected fetuses, we divided the prevalence rates for at-risk pregnancies by four, as the chance for the baby to be affected for recessively inherited disease is one in four (25%).

We further validated the assumptions by comparing the estimated prevalence per 100,000 pregnancies to the reported incidence rates for conditions covered (**Table S2**). While we could not validate the accuracy of alpha thalassemia estimates due to lack of reported data, numbers for other conditions were matched as closely as possible to the published data.

**Method S3: Calculation of screening costs per affected pregnancy**

It was determined that in the base scenario, 46 out of 110 affected fetuses would be identified per 100,000 pregnancies (**Table 2**). The total cost of screening for the base scenario is the sum of maternal carrier screen test ($694 per test × 100,000 pregnancies = $69.40M), the follow-up paternal testing (100,000 pregnancies × 16.1% carrier-positive rate × 41.5% paternal testing follow-up rate[2] × ($944 paternal testing and admin cost + 36% MFM referral rate × $1,236 MFM visit cost) = $9.2M), and the diagnostic testing cost for the at-risk couples (6,748 paternal testing run × 2.7% positive paternal testing rate × 50% diagnostic testing uptake × $710 diagnostic testing cost = $0.1M ). The total cost is therefore: $69.4M + $9.2M + $0.1M = $78.7M. To obtain the cost for identifying one affected pregnancy, we divided $78.7M by 46 (number of affected fetuses identified): $78.7M ÷ 46 = $1.73M.

In the reflex sgNIPT scenario, the total cost of screening is the sum of maternal carrier screen test ($69.4M) and the follow-up MFM visits (100,000 pregnancies × 0.7% abnormal sgNIPT result × 80% MFM referral rate × $1,236 MFM visit cost = $0.7M), and diagnostic testing cost (700 abnormal sgNIPT result × 50% diagnostic testing uptake × $710 diagnostic testing cost = $0.2M). The total cost is therefore: $69.4M + $0.7M + $0.3M = $70.4M. Reflex sgNIPT scenario identified 108 out of 110 affected pregnancies. Thus, the cost to identify one affected pregnancy is: $70.4M ÷ 108 = $0.65M.

The input assumptions are summarized in **Table 1** (model input assumptions), **Table 2** (number of affected fetuses identified), and **Table S3** (cost inputs). For the simplicity of the model, we assumed diagnostic testing uptake upon at-risk results is 50%, which is based on the estimate that the termination rates for these conditions range from 14% - 95% (**Table S7**).

**Method S4: SMA Treatment Cost Calculations**

***SMA types, life expectancies, and drug cost inputs***

SMA, with its phenotypic variation, are classified clinically into at least four types.[3,4] Type I is the most severe form, symptoms presenting at birth or in the first few months. Type II is intermediate and symptoms usually appear at ages of 7-18 months. Type III is mild and symptoms start to emerge from 18 months to early adulthood. Type IV (adult) is rare and symptoms do not emerge until second or third decade of life. Without treatment, Type I SMA children do not survive beyond two years of age. With the three treatments on the market, Onasemnogene abeparvovec (Zolgensma), Nusinersen (Spinraza), and risdiplam (Evrysdi), we assumed that the life expectancy for Type 1 SMA is extended to 15 years. Type 2 SMA life expectancy is not well known, but majority live into early adulthood.[5] With improved treatment options, we assumed that it is extended to 40. As the lifespan of Type 3 SMA is similar to the general population, we assumed it to be 70.

***Treatment choice difference between base scenario and reflex sgNIPT scenario***

To date, three major medications are available for SMA treatment: Onasemnogene abeparvovec (Zolgensma), Nusinersen (Spinraza) and the more recently FDA approved Risdiplam (Evrysdi). Between Zolgensma and Spinraza, Zolgensma has emerged as the therapy of choice for the majority of infants who are diagnosed before age of two.[6,7] In the simulated model, we assumed that 90% of SMA families under reflex sgNIPT flow would choose Zolgensma due to increased number of at-risk fetuses identified prenatally, where families can seek out diagnostic testing at birth and arrange the therapy early. In the base scenario, we assumed that 50% of families would choose Spinraza due to majority of the affected SMA babies being missed at the prenatal screening, which may delay the diagnosis and miss the window of treatment for Zolgensma. Due to the limited data availability, the use of Evrysdi is excluded from this simulation model.

For patients treated with Zolgensma, the lifetime treatment cost is assumed to be the cost of Zolgensma, i.e. $2.1M (**Table S4**). This is a simplified assumption that does not take into account of any related hospitalization costs and additional care costs. Thus, this is the conservative estimate of the total treatment cost.

For patients treated with Spinraza, we assumed the lifetime treatment cost is the cost of Spinraza per year (**Table S4**) multiplied by the life expectancy of each SMA type. We then applied the patient distribution by SMA type to obtain the weighted average. The resulted weighted average of SMA lifetime treatment cost is $8.5M for the base scenario and $3.4M for the reflex sgNIPT scenario (**Table S5**).

**Method S5: Calculation for decrease in lifetime treatment costs due to increased access to potential termination**

The decrease in lifetime treatment costs from increased access to potential termination per 100,000 pregnancies were calculated for each condition (cystic fibrosis, SMA, sickle cell disease/beta thalassemia, and alpha thalassemia). Decrease in lifetime treatment costs from termination for each condition = (# fetuses missed in base scenario; **Table S2**) × (% termination rate; **Table S7**) × (lifetime treatment cost; **Table S6**). Because termination rates vary considerably and data is limited, the minimum and maximum decrease in lifetime treatment costs (corresponding to 0% and 100% termination for each condition) were also calculated.

**Method S6: Pricing simulations for carrier screen with sgNIPT**

We took a value-based approach to simulating appropriate pricing for carrier screen with reflex sgNIPT (priced together) and reflex sgNIPT (priced separately), using two different unit costs.

1) Unit cost = cost to identify one affected fetus. The value of reflex sgNIPT is $1.73M (base cost to identify one affected fetus) - $0.65M (reflex sgNIPT cost to identify one affected fetus) = $1.08M per one affected fetus. The price of carrier screen with reflex sgNIPT with different value percentages passed to the price is calculated in the following way:

$$\text{Cost to identify one affected fetus} \\ = \big(\text{value of reflex sgNIPT}(\$1.08\text{M}) \times \text{percent value passed} \\ + \text{reflex sgNIPT cost per affected fetus } (\$0.65)\big)$$

$$\text{Total cost of screening and followup} \\ = \big(\text{cost to identify one affected fetus } \times \text{ affected fetuses detected } (108)\big)$$

$$\text{Price per test} = \text{total cost of screening and followup} \\ - \text{total cost of followup } (\$1\text{M}) \ / \ 100{,}000 \text{ pregnanices}$$

$$\text{Price per test if sgNIPT price separately} \\ = \text{total cost of screening and followup} \\ - \text{total sgNIPT cost of screening and followup } (\$70.4\text{M}) \\ /\text{number of carriers } (16{,}100)$$

2) Unit cost = total cost for 100,000 pregnancies. The value of reflex sgNIPT is $139.0M (base cost per 100,000 pregnancies) - $101.4M (reflex sgNIPT cost per 100,000 pregnancies) =

$37.6M per 100,000 pregnancies. The price of carrier screen with reflex sgNIPT is with different value percentages passed to the price is calculated in the following way:

$$\text{Screening and SMA cost per 100,000 pregnancies}$$
$$= \left(\text{value of reflex sgNIPT(\$37.6M)} \times \text{percent value passed} + \text{reflex sgNIPT cost per 100,000 pregnancies (\$101.4)}\right)$$

$$\text{Screening cost per 100,000 pregnancies}$$
$$= \text{ screening and SMA cost per 100,000 pregnancies} - \text{ sgNIPT SMA cost (\$31.1M)}$$

$$\text{Price per test} = \text{ [screening cost per 100,000 pregnancies} - \text{sgNIPT followup cost (\$1M)]} / \text{100,000 pregnancies}$$

$$\text{Price per test if sgNIPT price separately}$$
$$= \text{screening cost per 100,000 pregnancies} - \text{sgNIPT cost of screening and followup (\$70.4M)} / \text{number of carriers (16,100)}$$

**TABLE S1.** A summary of the demographic characteristics of the study population and carrier frequency input for the model analysis.

| Variables | Input Value |
| --- | --- |
| **U.S. ethnicity breakdown**[1] | |
| Northern European | 55.8% |
| African American | 13.9% |
| Hispanic | 20.8% |
| Asian | 7.2% |
| Other | 2.3% |
| **Carrier frequency** | |
| **Cystic fibrosis**[8] | |
| Northern European | 4.0% |
| African American | 1.6% |
| Hispanic | 1.7% |
| Asian | 1.1% |
| Other | 2.2% |
| **Spinal muscular atrophy**[9] | |
| Northern European | 2.1% |
| African American | 1.4% |
| Hispanic | 1.5% |
| Asian | 1.7% |
| Other | 1.9% |
| **Sickle cell disease/beta hemoglobinopathies**[10] | |

| | |
|---|---|
| Northern European | 0.6% |
| African American | 12.5% |
| Hispanic | 5.9% |
| Asian | 1.9% |
| Other | 2.0% |
| **Alpha-thalassemia**[10] | |
| Northern European | 2.3% |
| African American | 33.3% |
| Hispanic | 6.3% |
| Asian | 6.3% |
| Other | 6.3% |
| **% women who are carriers of one of the above disorders** | 16.1% |

**TABLE S2.** Estimated prevalence of affected fetus and reported incidence of the diseases.

| Variables | Estimated prevalence per 100,000 pregnancies | Reported incidence rate of affected births |
|---|---|---|
| **Total number of affected fetuses** | **109.9 (1 in 910)** | |
| Cystic fibrosis | 25.3 (1 in 3,950) | 1 in 3,900[11] |
| Spinal Muscular Atrophy | 8.8 (1 in 11,360) | 1 in 11,000[12] |
| Sickle cell disease/Beta-thalassemia | 73.6 (1 in 1,360) | 1 in 1540 for SCD only[13] |
| Alpha-thalassemia | 2.2 (1 in 45,450) | 1 in 10,000 in California; no national data[14] |

**TABLE S3.** Estimates of procedure costs.

| Variables | Input value |
|---|---|
| Maternal and paternal carrier screen cost | $694* |
| Carrier screen with reflex sgNIPT cost | $694* |
| Extra prenatal visit, counseling, and admin cost for paternal testing | $250** |
| Cost of MFM referral visit (perinatology consult, GC counseling, ultrasound) | $1,236*** |
| Cost of diagnostic testing (amniocentesis or CVS) | $730**** |

* Estimated cost based on CMS Clinical Laboratory Fee Schedule for cystic fibrosis (CPT 81220; $557) and spinal muscular atrophy (CPT 81329; $137) testing.

** We assumed one extra prenatal visit ($200) and two hours of extra admin work to arrange paternal testing ($50).

*** We assumed the average cost to be 200% of Maternal Fetal Medicine fee sheet 2018 published by Kaiser Permanente for its deductible plan members ($193.06 for MFM prenatal consult, $79 for genetic counseling, $345.58 for second trimester detailed scan).[15]

**** We used CMS 2021 Clinical Laboratory Fee Schedule for the cost of amniocentesis or CVS (CPT Codes 59000, 76946, 88235, 88267, 88280, 88291)

MFM, maternal fetal medicine; CVS, chorionic villus sampling.

**TABLE S4.** Assumptions for SMA treatment cost calculations.

| Variables | Input Value |
|---|---|
| SMA patient type distribution | |
| Type I (severe) | 58% |
| Type II (intermediate) | 29% |
| Type III (mild) | 13% |
| SMA life expectancy by Type | |
| Type I (severe) | 15 |
| Type II (intermediate) | 45 |
| Type III (mild) | 70 |
| SMA drug cost | |
| Zolgensma cost | $2,100,000 |
| Spinraza cost (initial dose) | $750,000 |
| Spinraza cost (ongoing dose) | $458,652 |

SMA, spinal muscular atrophy.

**TABLE S5.** Lifetime treatment costs of SMA.

| Variables | Value |
|---|---|
| Choice of treatment | |
| % SMA patients who choose Zolgensma in base scenario | 50% |
| % SMA patients who choose Zolgensma in reflex sgNIPT scenario | 90% |
| Lifetime treatment cost (weighted average across different SMA types) | |
| Base scenario | $ 8,499,707 |
| Reflex sgNIPT scenario | $ 3,350,807 |

SMA, spinal muscular atrophy.

**TABLE S6.** Lifetime treatment costs for disorders that are screened.

| Conditions | Life expectancy | Lifetime treatment cost ($MM) |
|---|---|---|
| CF | 44-47 years[16] | $ 7.03[17] |
| SMA | <2 years to adulthood | $ 8.50[*] |
| Hemoglobinopathies** | 42-47 years[18] | $ 0.46[19] |
| Weighted average | N/A | $ 2.62 |

* Detailed calculation is described in **Method S4**.

** Hemoglobinopathies have a wide range of clinical outcomes. For alpha thalassemia, the clinical outcomes can range from stillbirth (alpha thalassemia major) to mild symptoms that do not alter life expectancy (Hemoglobin H). Because the treatment cost data is limited for hemoglobinopathies other than sickle cell disease, the cost estimate is based on a study on sickle cell disease.

CF, cystic fibrosis; SMA, spinal muscular atrophy.

**TABLE S7**. Sensitivity Analysis for Paternal Follow-up Testing Rate

| | Paternal Follow-up Rate | | |
|---|---|---|---|
| | **Medium** | **Low** | **High** |
| **Costs** | **(42%)** | **(20%)** | **(80%)** |
| *Base scenario* | | | |
| Follow-up cost per 100,000 pregnancies ($MM) | $78.71 | $73.89 | $87.35 |
| # Affected fetuses identified | 46 | 22 | 88 |
| Cost to identify one affected fetus ($MM) | $1.73 | $3.36 | $0.99 |
| *Reflex sgNIPT scenario* | | | |
| Follow-up cost per 100,000 pregnancies ($MM) | $70.36 | ” | ” |
| # Affected fetuses identified | 108 | ” | ” |
| Cost to identify one affected fetus ($MM) | $0.65 | ” | ” |
| Follow-up Cost Savings per 100,000 pregnancies ($MM) | $8.35 | $3.52 | $16.99 |
| Follow-up Cost Savings per Affected Fetus (fold decrease) | 2.65 | 5.17 | 1.53 |

**TABLE S8**. Decrease in Lifetime Treatment Costs.

| | Condition | | | | |
|---|---|---|---|---|---|
| **Item** | **CF** | **SMA** | **SCD/Beta-thal** | **Alpha-thal** | **Total** |
| **TOP Decrease in Lifetime Treatment Costs ($MM)** | **$95.34** | **$31.44** | **$17.91** | **$0.08** | **$145.38** |
| Range 0%-100% TOP ($MM) | $0 to $101.42 | $0 to $34.29 | $0 to $19.30 | $0 to $0.58 | $0 to $155.59 |
| *Fetuses affected* | 25.3 | 8.8 | 73.6 | 2.2 | 109.90 |
| *Fetuses missed in base scenario* | 14.4 | 5.0 | 42.0 | 1.3 | 62.6 |
| *Lifetime treatment cost ($MM)* | $7.03 | $6.84 | $0.46 | $0.46 | - |
| *TOP rate (%)* | 94%[20] | 92%[21] | 93%[22–24] | 14%* | - |

CF, cystic fibrosis; SMA, spinal muscular atrophy; SCD, sickle cell disease; TOP, termination of pregnancy

*Internal calculation based on the assumption of 90% miscarriage/stillbirth/abortion rate for alpha thalassemia major (<5% of cases) and Hb CS (10%), and 0% termination for HbH (>85%).
